# Oral CBD treatment is associated with an anti-inflammatory gene expression signature in myeloid cells of people living with HIV

**DOI:** 10.1101/2023.02.24.23285761

**Authors:** Simone Marini, Amanda Huber, Melanie N. Cash, Marco Salemi, Robert L Cook, Paul Borsa, Carla N. Mavian

## Abstract

HIV-related comorbidities appear to be related to chronic inflammation, a condition characterizing people living with HIV (PLWH). Prior work indicates that cannabidiol (CBD) might reduce inflammation; however, the genetics underpinning of this effect are not well investigated. Our main objective is to detect gene expression alterations in human peripheral blood mononuclear cells (PBMCs) from PLWH after at least one month of CBD treatment.

We analyze ∼41,000 PBMCs from three PLWH at baseline and after CBD treatment (27-60 days). We obtained a coherent signature, characterized by an anti-inflammatory activity, of differentially expressed genes in myeloid cells.

Our study shows how CBD is associated with alterations of gene expression in myeloid cells after CBD treatment.

## Introduction

Despite the advent of combined antiretroviral therapy (cART), residual HIV replication persists and is associated with low-level immune activation driving chronic inflammation, one of the main HIV-related complications. PLWH experience non-AIDS-defining illnesses associated with age that are typical of older patients, such as presence of neurocognitive disorders, metabolic syndrome, bone abnormality, non-HIV-associated cancers, and neurocognitive disorders.^1^ Chronic inflammation is a mechanism broadly involved in a plethora of diseases affecting aging patients. Experts explicitly name this condition as *inflammageing*: Inflammageing is a pivotal risk factor for dementia, chronic kidney disease, diabetes mellitus, cancer, depression, cardiovascular diseases, and sarcopenia^2^.

While the main HIV reservoir is known to be in resting memory CD4+ T-cells, several studies have shown that a fraction of viral variants, detected either in residual viremia during therapy or at rebound after therapy interruption, is genetically distinct from virus present in T-cell subsets.^3-5^ Myeloid cells, particularly monocytes and macrophages, have been described as active reservoirs driving persistent low-level HIV expression and contributing to viral replication during rebound.^6-10^

Monocytes are chronically activated during HIV infection, and are major mediators for the development of comorbidities related to inflammageing, and specifically cardiovascular diseases, neurocognitive disorder, and aging of the innate immune system.^11,12^ Residual HIV viremia is likely a source of monocyte activation as HIV proteins, such as p17, induce the production of proinflammatory molecules by activated monocytes.^13^ Inflammatory mediators produced by monocytes, but not T-cell activation, predict low levels of chronic immune activation that persist during cART treatment, a process named serious non-AIDS-defining illnesses associated with age during viral suppression on cART,^14,15,16^ highlighting the important role of monocyte activation during cART-treated HIV infection.

*Cannabis sativa* is widely used for medical purposes and has anti-inflammatory activity. Cannabis sativa and its cannabinoid derivatives, Δ9-tetrahydrocannabinol (THC) and cannabidiol (CBD), are largely used by people living with HIV (PLWH) to stimulate appetite, prevent weight loss (FDA approved Marinol or Dronabinol), or manage chronic pain. CBD does not have psychoactive effects and CBD products are available over the counter in all the United States^17,18^. CBD is a compound investigated for its beneficial effect on inflammation;^19-20^ but despite its widespread use, the impact of CBD on health outcomes in PLWH remains understudied.^22^ Here we investigate the hypothesis that CBD is associated with a reduction of the expression of proinflammatory genes in PBMCs of PLWH via single-cell RNA sequencing (scRNAseq).

Our results show a downregulated gene expression signature associated with CBD in myeloid cells. Importantly, the great majority of these genes are known to be inflammation promoting.

## Results

We sequenced six blood samples from three PLWH (IND1, IND2, IND3): for each individual, we included a baseline (pre-CBD treatment) and a post-CBD treatment (27-60 days, ∼67mg/day).

### We identified eight distinct cell populations based on their expression profiles

We used the Seurat package^23^ for downstream analysis (Figure 1, A-D), applied after obtaining the results from 10x Genomics Cell Ranger 3.0.0. Details of the procedure are reported in the Methods. After quality control, we obtained 30,339 cells expressing 41,198 genes. Unsupervised clustering revealed 10 cell clusters (Figure 1, A). Per-individual projection confirms that cells from each individual are present in all the clusters (Figure 1, B).

**Figure 1.**
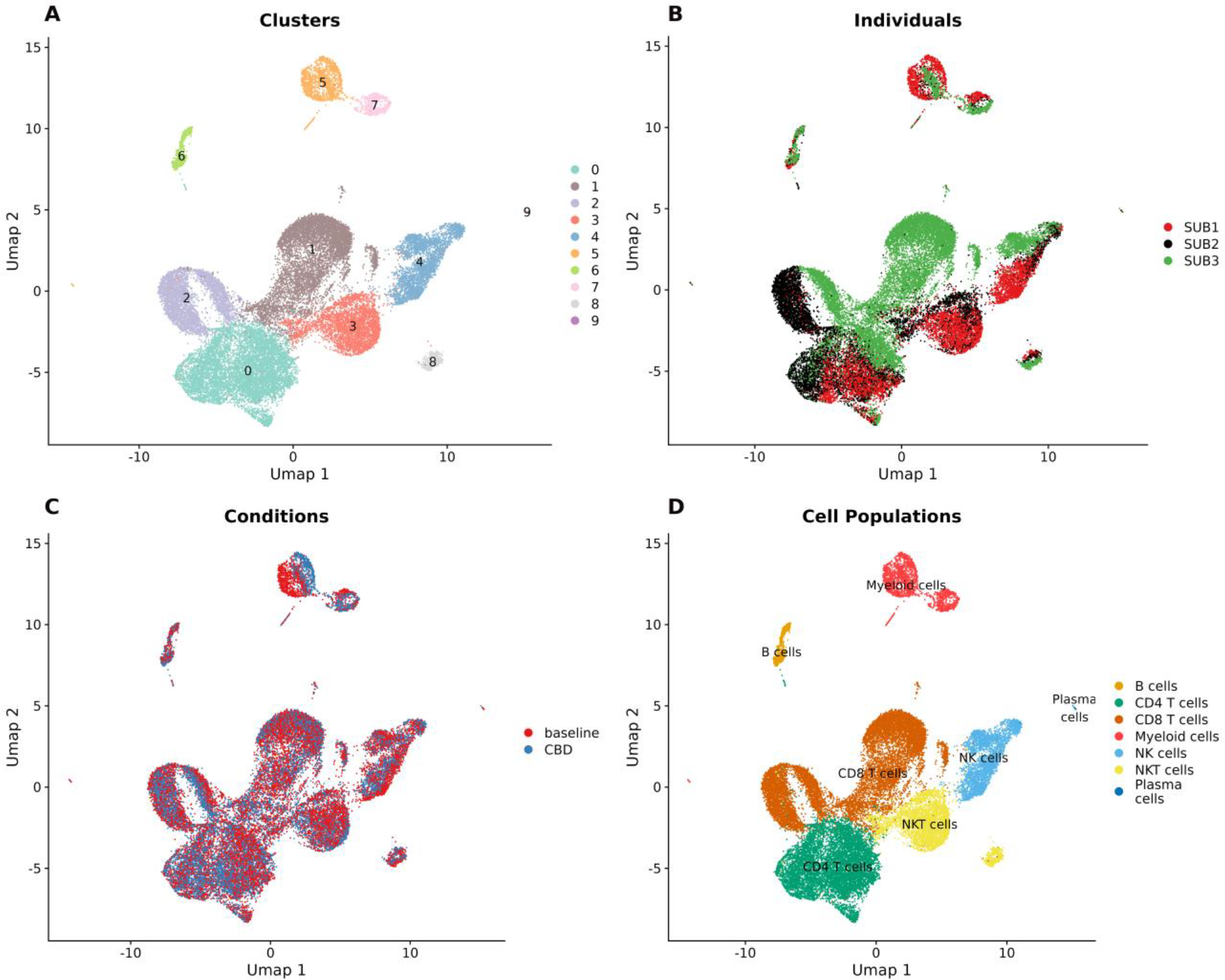
UMAP projection of the dataset. (A) Unsupervised clustering finds 10 cell clusters, numbered 0 to 9. (B, C) Per-individual and per-condition colored projections. (D) Cell populations labeled based on overexpressed genes in the clusters.

We labeled the clusters based on specific genes that are characterizing cell populations (Figure 2). We obtained seven cell populations (Figure 1, D): *CD4* T cells (cluster 0; 8,622 cells expressing *IL-7R, LEF1, CD28*, but not *CD8A*); *CD8* T cells (clusters 1 and 2; 11,303 cells expressing *CD8A*); NKT cells (clusters 3 and 8; 4,279 cells expressing *NKG7, KLRD1, KLRG1*, and *CD8A*); NK cells (cluster 4; 3,223 cells expressing *NKG7, KLRD1, KLRG1*, but not *CD8A*); Myeloid cells, including monocytes (clusters 5 and 7; 2,277 cells expressing *LYZ, CD14, S100A8*, and *S100A9*); B cells (cluster 6; 595 cells expressing *CD79A* and *CD79B*); and plasmablasts/plasma cells (cluster 9; 40 cells, expressing *JCHAIN, IGHG1, IGHA1, IGHA2*).

**Figure 2.**
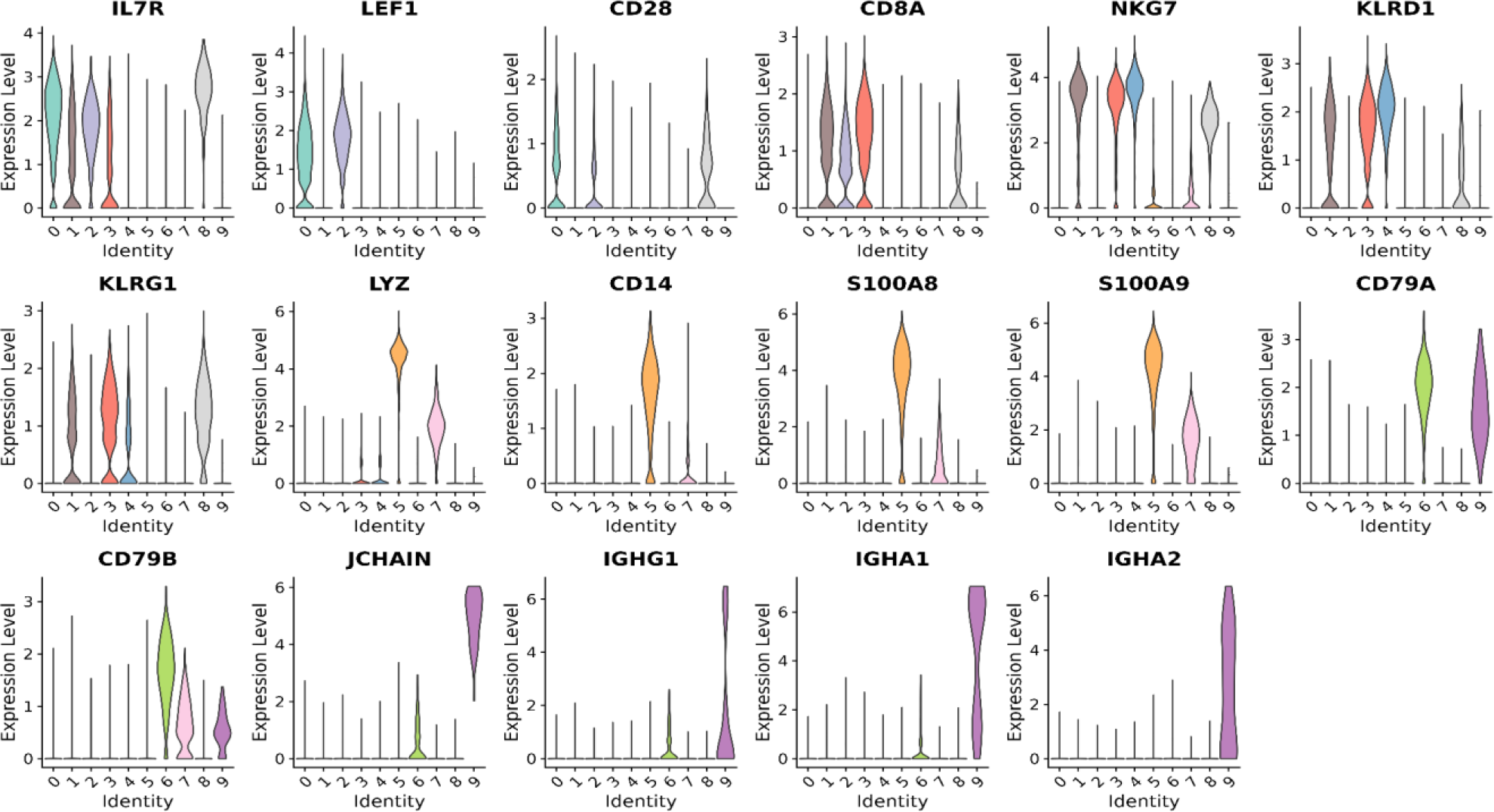
Gene expression violin plots for genes of interest to label cell clusters into populations: Cluster 0, CD4 T cells; clusters 1 and 2, CD8 T cells; clusters 3 and 8, NKT cells; cluster 4, NK cells; clusters 5 and 7, myeloid cells, including monocytes; cluster 6, B cells; and cluster 9, plasmablasts/plasma cells.

### Myeloid cells carry a CBD-associated anti-inflammatory gene expression signature

We identified differentially expressed genes (DEGs) in myeloid cells, reported in Table 1. Since the number of myeloid cells is different for the three individuals, we provided DEGs for the cell population considering all the individuals together, and for each distinct individual. We consider as DEGs the six genes that are differentially expressed both overall (i.e., consider all the myeloid cells) and in at least two of three PLWH (i.e., considering only the cell from a single person). All the DEGs are under-expressed after CBD treatment. Five out of six genes show a positive role in the inflammation process: *CXCL8* (interleukin-8) is a member of the chemokine family involved in cell recruitment and activation under inflammatory conditions;^24^ *EREG* is a proinflammatory gene with a role in tissue healing and vascularization;^25^ *JUN* and *FOS* are two genes expressed in monocytes and macrophages involved in promoting inflammation,^26,27^ and both are included as markers in a recently general inflammatory macrophage activation phenotype encompassing responses to HIV, sepsis, and SARS-CoV-2^28^ *CCL3* is a proinflammatory gene and one of the major HIV-suppressive factors produced by CD8+ T-cells and monocytes.^28,29^ Other proinflammatory genes appear among the top under-expressed genes (Table 1): *GPR183* plays a role in attracting macrophages during viral inflammation;^30^ *MIR23AHG* is a long non-coding RNA gene promoting inflammation polarization in macrophages^31^ and associated to chronic inflammation in ulcerative colitis;^32^ *TIPARP* is an antiviral gene, with a specific anti-HIV-1 effect in monocytes;^33^ *NAMPT* is a gene involved in the inflammatory responses activation with cytokine-like behavior;^34^ *IL1B* is a proinflammatory cytokine;^35-36^ and *KLF6* is known to regulate macrophage polarization towards the proinflammatory phenotype.^37^ In summary, five out of six DEGs and several of the top under-expressed genes are markedly involved in inflammatory processes of myeloid cells. Taken together, this concerted underexpression can therefore be interpreted as an anti-inflammatory signature associated with the CBD treatment.

**Table 1.** Top under-expressed genes in myeloid cells (log2FC < -0.9, considering cells from all the individuals). **Bold**: DEGs; underlined: genes of interest; log2FC = base-2 logarithm of the fold change (CBD versus baseline); Fraction baseline = fraction of cells expressing the gene at baseline; Fraction CBD = fraction of cells expressing the gene after CBD treatment; P-value (adj.) = FDR adjusted p-value; values reported as * imply a logFC within the [-0.1, 0.1] interval.

| *Gene* | *log2FC (all)* | *log2FC IND1* | *log2FC IND2* | *log2FC IND3* | *Fraction Baseline (all)* | *Fraction CBD (all)* | *P-value (adj., all)* |
| --- | --- | --- | --- | --- | --- | --- | --- |
| **CXCL8** | -1.78 | -2.03 | -2.03 | * | 0.558 | 0.344 | 0 |
| **EREG** | -1.72 | -1.87 | -1.92 | -0.83 | 0.304 | 0.149 | 0 |
| **PLEKHG2** | -1.53 | -1.92 | -1.62 | -0.63 | 0.571 | 0.21 | 0 |
| G0S2 | -1.24 | -1.76 | -0.14 | 0.15 | 0.21 | 0.21 | 1.27E-253 |
| **JUN** | -1.23 | -1.38 | -0.83 | -1.07 | 0.899 | 0.732 | 0 |
| **FOS** | -1.15 | -1.62 | -1.16 | -0.35 | 0.998 | 0.985 | 0 |
| <u>GPR183</u> | -1.08 | -1.42 | -0.69 | -0.39 | 0.427 | 0.21 | 1.82E-219 |
| <u>MIR23AHG</u> | -1.06 | -1.34 | -0.96 | -0.45 | 0.831 | 0.918 | 0 |
| **CCL3** | -1.04 | -1.32 | -1.38 | -0.39 | 0.366 | 0.174 | 6.62E-184 |
| <u>TIPARP</u> | -1.04 | -1.35 | -0.88 | -0.35 | 0.717 | 0.512 | 8.33E-275 |
| ABCA1 | -1.02 | -1.46 | -0.43 | -0.35 | 0.646 | 0.418 | 1.6e-254 |
| ENSG00000287124 | -0.99 | -1.06 | -0.77 | -0.73 | 0.645 | 0.501 | 1.33E-241 |
| <u>NAMPT</u> | -0.98 | -1.52 | -0.47 | * | 0.986 | 0.989 | 0 |
| ENSG00000253736 | -0.97 | -1.03 | -1.01 | -0.84 | 0.926 | 0.793 | 1.85E-301 |
| <u>IL1B</u> | -0.96 | -0.94 | -1.76 | -0.43 | 0.536 | 0.364 | 1.08E-269 |
| <u>KLF6</u> | -0.96 | -1.12 | -1.01 | -0.46 | 0.984 | 0.971 | 0 |
| ANKRD28 | -0.91 | -0.97 | -1.52 | -0.28 | 0.601 | 0.402 | 1.83E-171 |

### STRING analysis indicates the DEG signature includes a protein interaction module

Using our DEGs as input in the STRING database, a collection of known protein interactions,^38^ with highest stringency for the interaction score, we obtained a structured protein interaction module centered on *JUN* and including five out of six genes (p-value 0.0001, Figure 3A). We proceeded with a DEG pathway analysis by adding the highest-confidence STRING gene interactions to the DEGs, and obtaining a network of 56 nodes (Figure 3B). The pathway analysis of the network confirmed its proinflammatory nature, with focus on Mitogen-activated protein kinase (MAPK), Cytokine, and Interleukine pathways. Among the top 20 Reactome^39^ terms (ordered by p-value) we find Signaling by Interleukines (p-value 7.5e-23); Cytokine Signaling in Immune system (p-value 2e-21); Immune System (p-value 2e-17); Interleukine-10 signaling (p-value 2.1e-15); Interleukin-4 and Interleukin-13 signaling (p-value 4.7e-15); NGF-stimulated transcription (p-value 2.4e-12); Activation of AP-1 family of transcription factors (p-value 4.3e-12); Signaling by NTRK1 (TRKA) (p-value 1e-11); MAPK targets/Nuclear events mediated by MAP kinases (p-value 2.5e-11); Signaling by Receptor Tyrosine Kinases (p-value 1.15e-9); Toll Like Receptor 3 (TRL3) Cascade (1.2e-9); Chemokine receptors bind chemokines (1.2e-9); and TRIF(TICAM1)-mediated TLR4 signaling(1.5e-9). Supporting the above data suggesting a CBD-associated anti-inflammatory signature in myeloid cells, it is known that IL-4 and IL-13 are cytokines known to polarize myeloid cells towards an M2, anti-inflammatory phenotype^40^. Further, IL-10, an anti-inflammatory molecule, can polarize myeloid cells to an M2c anti-inflammatory phenotype, and M2 anti-inflammatory macrophages are known to produce this molecule^40,41^. All the retrieved pathways are attached in Supplementary Table 1.

**Figure 3.**
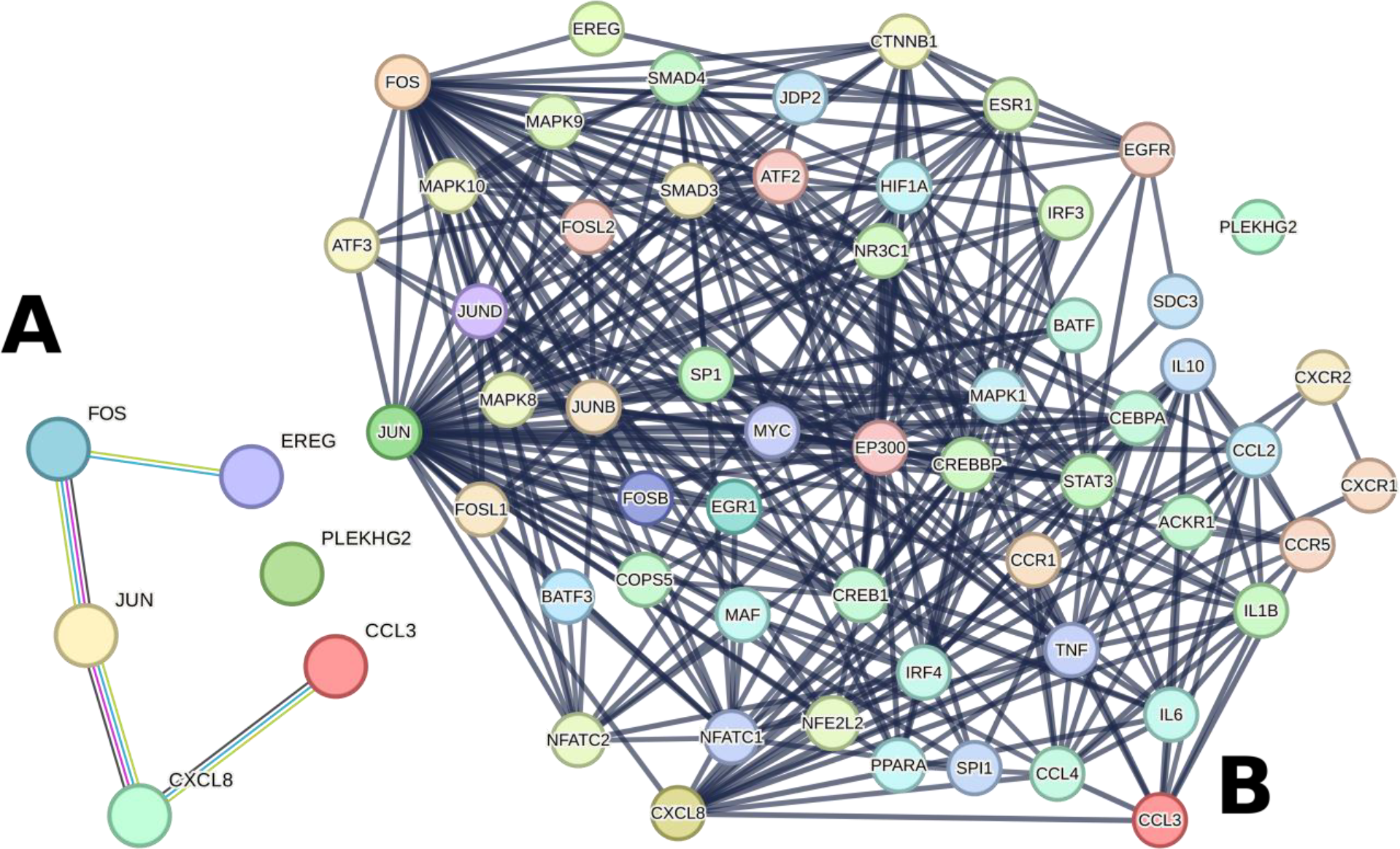
String analysis. (A) The coherent nature of the anti-inflammatory protein module (DEGs in CBD versus baseline) is confirmed as five out of six DEG show interactions in the STRING database analysis, most stringent interaction threshold, p-value 0.0001. (B) The gene module expanded with its first interactors.

### GO-based scoring and serology suggest a general anti-inflammatory shift in myeloid cell gene expression

To further confirm the anti-inflammatory shift in myeloid cells, we designed an inflammation score based on the gene expression of all the genes expressed in our dataset that are listed under the Gene Ontology term *Positive regulation of inflammatory response* (GO:0050729). The score decreases 12% after CBD in myeloid cells, from a median of 0.225 at baseline to a median of 0.198 (p-value = 9.538e-08, Kolmogorov-Smirnov test). This decrement is conserved by analyzing the score distribution per individual, as depicted in Figure 4, i.e., in IND1, from 0.169 to 0.157, p-value = 0.004226; in IND2 from 0.147 to 0.05, p-value = 6.741e-08; and in IND3 from 0.13 to 0.102, p-value = 3.524e-05.

**Figure 4.**
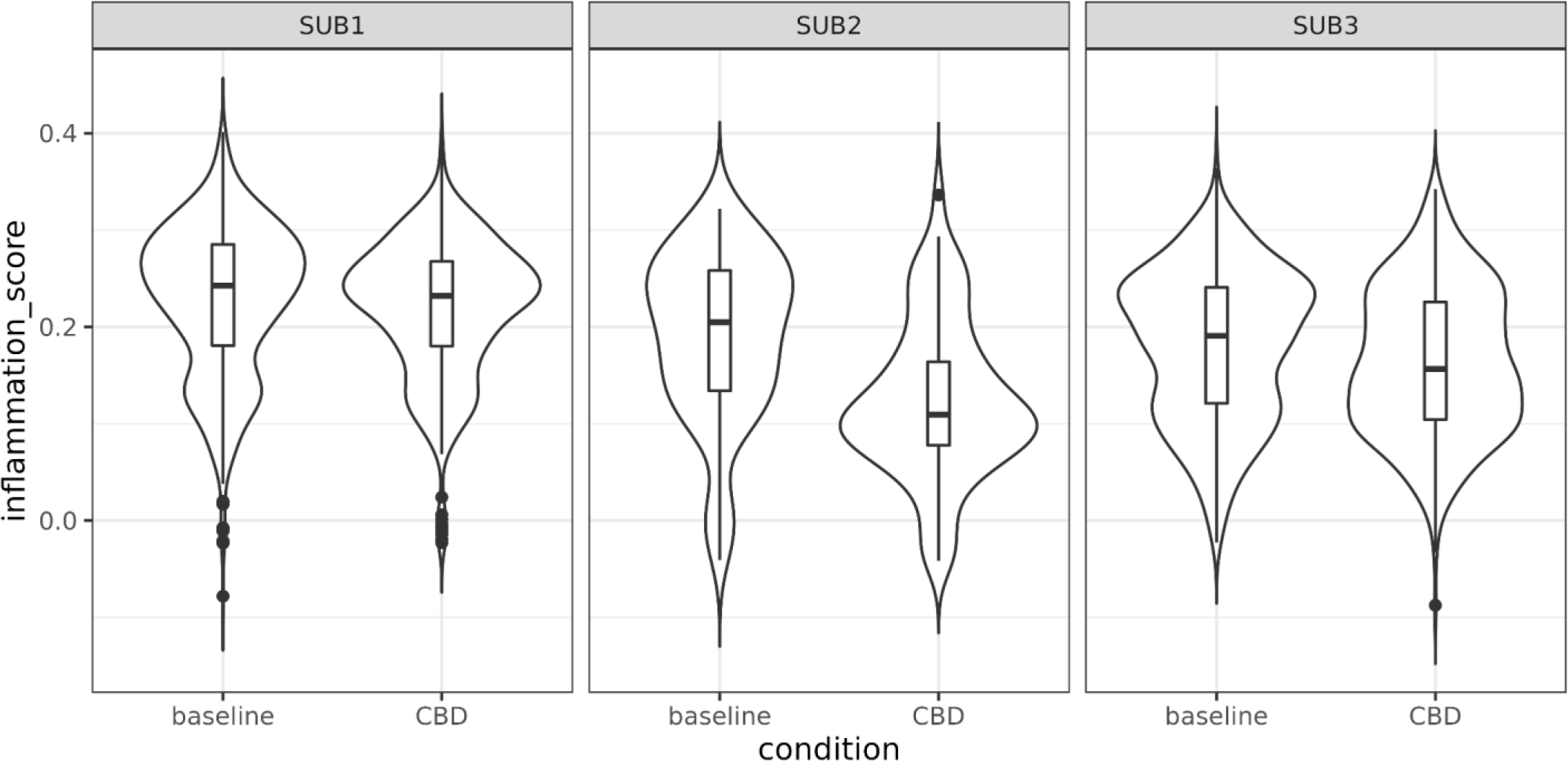
Violin and box plots of the inflammation score, per individual. The inflammation score is calculated with the genes of GO:0050729 (Positive regulation of inflammatory response). CBD score distributions are significantly lower in all individuals with p-values of 0.004226, 6.741e-08, and 3.524e-05 for IND1, IND2, and IND3 respectively.

The general reduction in inflammation is also indicated serologically. More specifically, the decline in c-reactive protein (CRP) measure (baseline versus post-CBD treatment) was 10.25 and 8.2 for IND1, 1.4 and 0.79 for IND2, 2.56 and 0.88 for IND3. The decline in erythrocyte sedimentation rate 32 and 30 in IND1, 13 and 10 in IND2, 12 and 4 in IND3.

### Inference of cell-cell communication variation

We used CellChat^42^ to analyze the variation of cell-cell communication between baseline and CBD conditions (Figure 5). The most affected interactions involve myeloid cells, with decreased interactions from CD8 T cells, B cells, CD4 T cells, NK cells, and with themselves; and with an increase of signals directed towards NKT cells.

**Figure 5.**
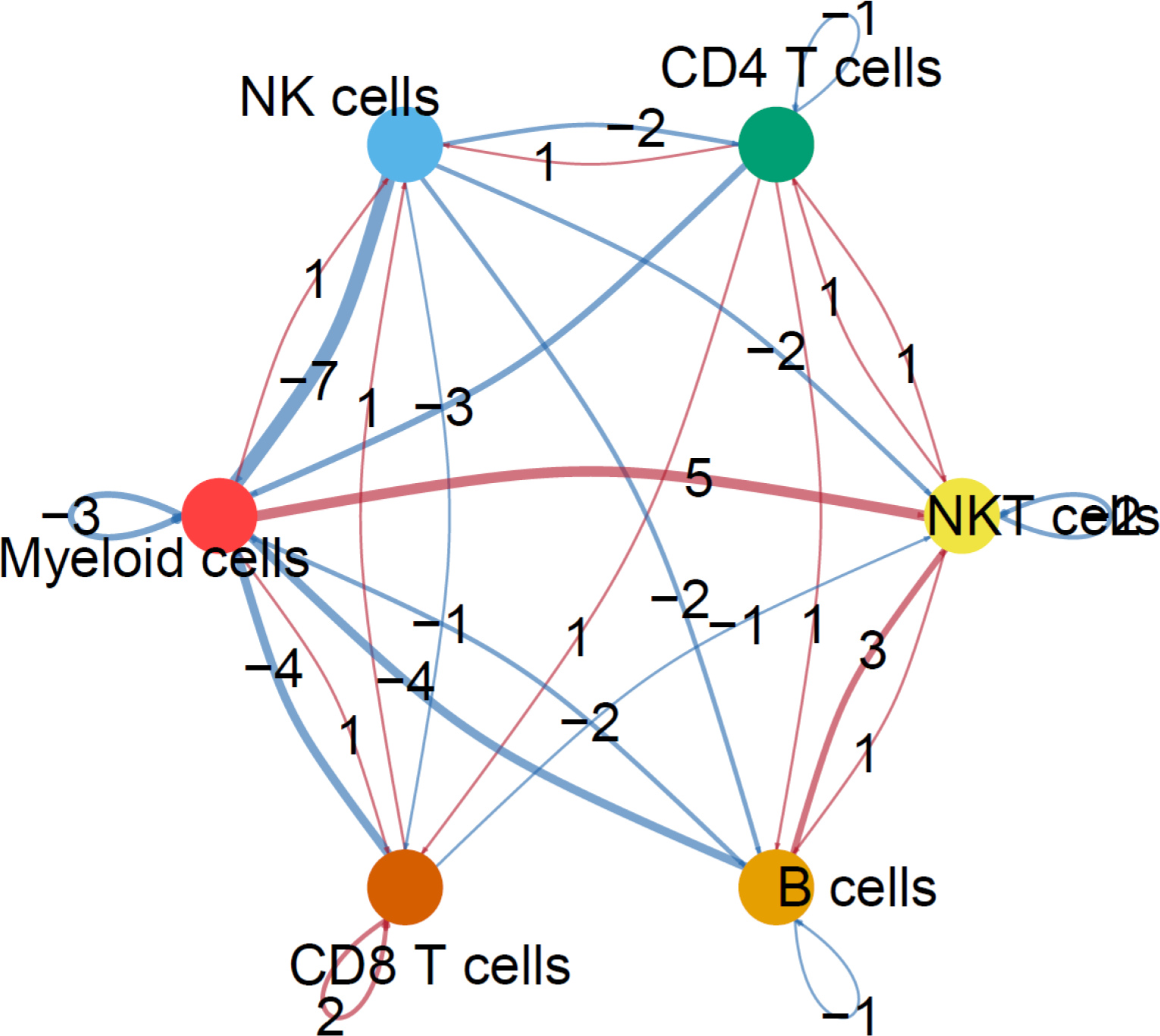
Cell-cell communication inferred by cell population.

### Differentially expressed genes in other cell populations

Generally, non-myeloid cell populations did not show differentially expressed genes, with the exception of B cells, where we found *GNLY* under-expressed after CBD. *GNLY* is a cytotoxic gene that is typically expressed in NK and T cells^41^. *GNLY* has shown the ability to promote the differentiation of monocytes into dendritic cells, and is known for its marked inflammatory role.^43,44^ Its decreased expression in the B cell population is in line with a general shift towards a less proinflammatory gene expression profile.

## Discussion and Conclusions

In this study, we measured gene expression at the single-cell level in PBMC samples obtained from three PLWH before (baseline) and after 27 to 60 days of CBD oil treatment. Our data indicate that CBD treatment is associated with decreased expression of a proinflammatory myeloid cell gene signature in PLWH on cART, enriched for specific inflammation pathways including MAPKs, cytokines, and interleukines. The CBD dosage of this study is within the dosage recommended by the CBD oil manufacturer, and is significantly lower than the one administered in other CBD clinical studies addressing, for example, psychosis or seizures.^46^ Of note, the CBD oil used in this study is hemp-derived and therefore different from purified synthetic CBD such as Epidiolex.

This study does not come without limitations: We processed data for individuals with different lengths of CBD exposure; and IND3 did not follow the CBD regimen as diligently as IND1 and IND2. This seems to be reflected in the gene expression pattern (Table 1), where IND1 and IND2 show more marked and uniform effects. However, the gene expression variation in IND3 seems to be in line with the other individuals, with 14 out of the top 17 under-expressed genes following the same trends. Furthermore, the anti-inflammatory shift in myeloid cell gene expression in IND3 is also supported by the GO-based score (Figure 4) and the reduction in CRP and erythrocyte sedimentation. The extracted gene module is highly consistent with a diminished expression of proinflammatory genes and pathways; however, we did not find any specific connection between PLEKHG2 (one of the DEGs) and the inflammatory phenomenon in literature. Consistent with this, PLEKGH2 does not share interactions with the extended network of first-intreactors of the proinflammatory gene module (Figure 3B), even if we change the STRING threshold from 0.9 (highest confidence) to 0.15 (low confidence). A possible relation could consist of PLEKGH2 to be involved in monocyte migration, as PLEKHO2, another gene with a plekstrine homology binding site, has been shown to be involved in this process^46^. Further investigations are needed to confirm and understand if PLEKHG2 plays a role in the inflammatory pathways, and whether its significantly decreased expression is associated with CBD.

Another limitation of this work is that, by limiting our age range, we did not include older adults (65+). As the use of CBD and cannabis sativa for medical purposes by older adults is increasing^47^, future studies should also focus on this population.

While this study helps understand the genetic underpinning of CBD anti-inflammatory action, and the potential of its anti-inflammatory effects, the molecular chain leading to inflammation reduction remains to be unveiled. In future studies, we aim to target the extracted gene signature to better characterize it, along with its molecular interactors and a measure of HIV viral load. As oral CBD has very low bioavailability, in future studies we plan to measure serum levels of CBD and other metabolites, and integrate the findings with the gene expression analysis in order to better understand the molecular CBD machinery beyond the RNA level.

## Methods

### Recruitment

All individuals were recruited according to a UF IRB-approved protocol and informed consent. *Main inclusion criteria included*: be older than 21 and younger than 60; be HIV+ and under antiretroviral therapy for 5+ years; report suppressed viral load in their most recent check-up; tests negative for THC; and be practicing birth control. Main exclusion criteria included: be suffering from conditions or taking medications that may impair the immune response; be pregnant; be a current marijuana or CBD user; be taking anti-seizure medicines; having any history of seizure disorder or head trauma; hepatic/liver diseases or renal diseases, such as cirrhosis or multicystic dysplastic kidney; or cardiovascular disease, such as coronary heart cerebrovascular diseases. We excluded all serious comorbidities. Eligibility for the study was confirmed by a physician.

### CBD treatment

CBD oil was provided by SunFlora, Inc, 411 19th Street South, St. Petersburg, FL 33712. SunFlora is a third-party tested hemp-derived product manufacturing company. The research team modified an existing FDA IND from SunFlora for this study. The hemp-extract (SunMed™ Hemp Supplement) is a full spectrum CBD-rich formulation (∼4-7%) containing very low levels of THC (<0.3%), along with all naturally occurring cannabinoids, terpenes, and essential oils from the plant extract. Flavoring agents have been added to ensure palatability. The tincture solution is provided in a 30mL bottle (2000mg CBD). Each individual was provided two bottles, each with a syringe dropper with instructions to administer the solution under the tongue twice per day (BID), 8-12 hours apart (morning and evening) with food at the prescribed daily dosage (1ml or ∼67mg of CBD/day). The study protocol was approved by University of Florida’s IRB. All methods were performed in accordance with the relevant guidelines and regulations.

### Sample collection

We sequenced six blood samples from three PLWH (IND 1-3): 2 females and one male; mean age 44.3±9.5; reported cART treatments were valaciclovir, dolutegravir/lamivudine, bictegravir/emtricitabine/tenofovir alafenamide, and darunavir/cobicistat. All individuals reported a viral load of less than 200 copies of HIV per milliliter of blood at their last check-up. Two samples are included in this study for each individual: a baseline (pre-CBD treatment) and a post-CBD treatment. The post-CBD treatment sample was taken after 27 days of treatment for IND1, with ∼8% missed doses; after 70 days for IND2 (total 60 days of treatment), with ∼6% missed doses; and after 29 days for IND3 with ∼60% missed doses. The planned treatment was 60 days, however, it was not possible to consider samples after the first 30 days from IND1 and IND3. Sedimentation Rate and CRP were obtained through the lab services of the UF Health Shands Hospital.

### Single-cell extraction and sequencing

*PBMC extraction*: Blood was collected in vacutainer EDTA tubes and centrifuged within 30 minutes of collection at 200 x g for 10 minutes to separate plasma and cellular fraction. Blood fraction was diluted with an equal amount of wash buffer (Phosphate buffered saline [137 mM NaCl, 2.7 mM KCl, 8 mM Na2HPO4, and 2 mM KH2PO4]; 1mM EDTA) and mixed by inversion 5 times. Diluted blood was carefully layered over Lymphoprep Separation Media (Cellgro, Lincoln, NE) by gravity release. Tubes were centrifuged at 400 x g for 30 minutes in a swinging bucket centrifuge with brake off. The PBMC layer was collected, and cells were washed twice with wash buffer upon centrifugation at 400 x g for 10 minutes at room temperature with brake on. Next, cells were subjected to a lower speed wash at 200 x g for 10 minutes. Cells were filtered in a 70μM strainer (Fisher, Hampton, NH), counted and frozen down using CryoStor Freezing Medium (Stem Cell Technologies, Vancouver, BC, Canada) at ∼15 million PBMCs per patient until they could be used for single-cell sequencing. *Sample preparation for single-cell RNA sequencing*: Cryopreserved human peripheral blood mononuclear cells (PBMCs) were prepared for 10x Genomics® Single Cell Sequencing according to the manufacturer’s protocol (Pleasanton, CA) with a few modifications. Briefly, the cells were quickly thawed at 37°C following dropwise addition of warm complete medium (10% FBS in DMEM (Fisher)). Following centrifugation, cells were washed once again with complete medium before being subjected to dead cell removal using the Dead Cell Removal Kit (Miltenyi Biotech, Waltham, MA) according to the manufacturer’s protocol. Next, the cells were pelleted and washed with PBS, 0.04% BSA (400 μg/mL) twice before filtering through a Flowmi™ Tip Strainer (Fisher) and counted to resuspend the cells at 1200 cells per μL for viability testing and library preparation. *Library construction and sequencing:* The 10x Chromium Controller and Single Cell 3′ Library & Gel Bead Kit v3 (10X Genomics) were used to construct barcoded single-cell RNA-Seq libraries following the manufacturer’s user guide (10X Genomics). Briefly, approximately 10,000 single cells at 90% or greater viability are loaded onto the Chromium Controller. Single cells are captured into Gel Bead-In Emulsions (GEM) where cell lysis and cDNA synthesis occur. All cDNAs produced from a single cell will contain a unique DNA molecular barcode. Following demulsification, mRNA-derived are recovered using SPRIselect reagent (Beckman). Library construction was performed by fragmentation, end repair, A-tailing, adaptor ligation, and sample index PCR, to which Illumina sequencing barcodes were added. After Ampure Beads clean up, libraries were quantified by using Agilent TapeSation 4200 and qPCR quantification with NEB Library Quanti Kit for Illumina (NEB). *Sequencing*: The libraries were sequenced on an Illumina NovaSeq 6000 instrument using the S4 Reagent Kit v1.5, and aiming for 85,000 reads per cell. We obtained 6,202.5 ± 1,604.09 cells per sample, and 572,746,422 ± 183,985,985 reads per sample (Table 2). Count matrices were obtained with 10x Genomics Cell Ranger Single Cell Software Suite. Library preparation and scRNASeq were performed at the UF Interdisciplinary Center for Biotechnology Research (ICBR), University of Florida (UF). All quality control measures were in the normal range.

**Table 2.**
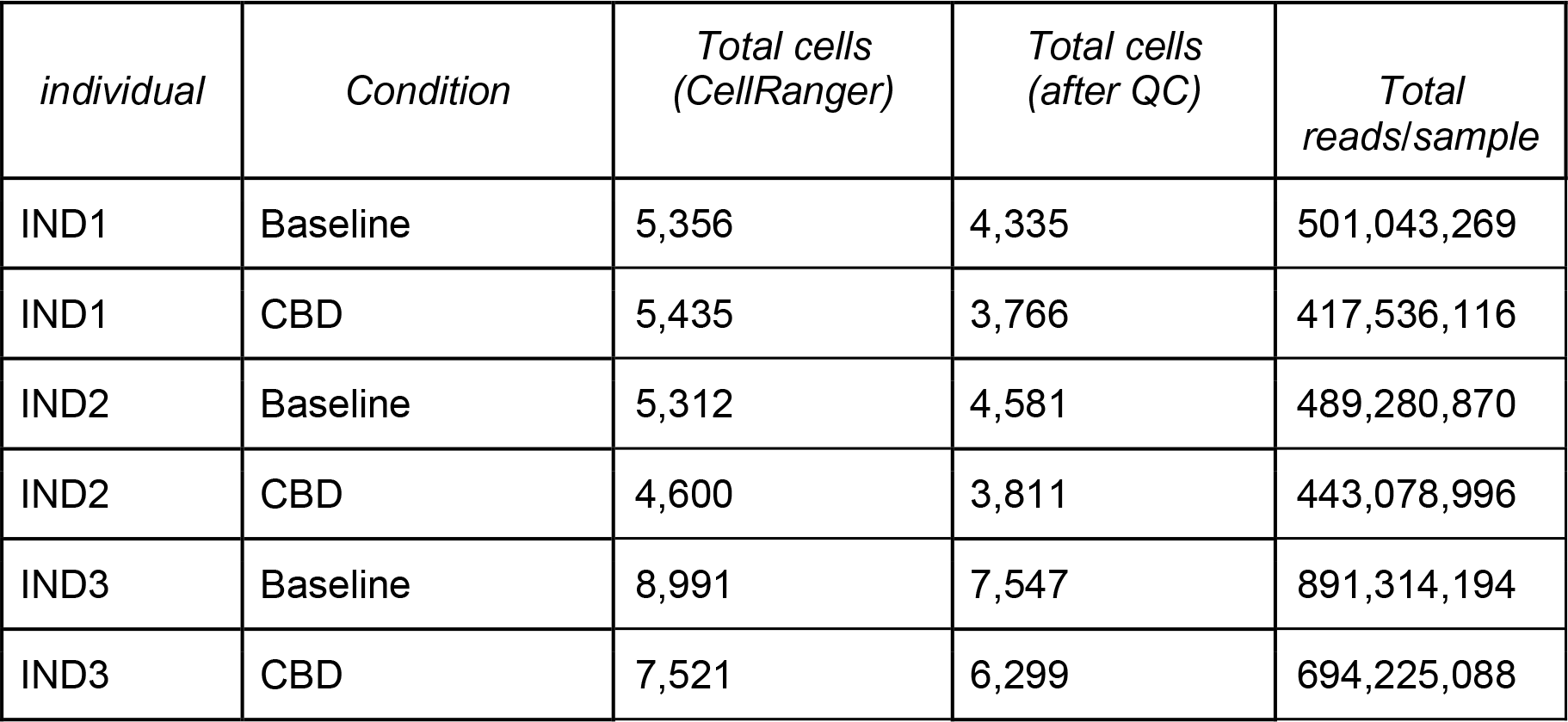
Total cells and reads per sample.

### Single-cell downstream analysis

The analysis is based on Seurat v4.2.0^23^. Where not indicated, parameters were used at their default values. Briefly, we merged the six samples and proceeded with quality control based on: number of different genes expressed per cell, counts per cell, and fraction of mitochondrial content per cell. We based our downstream analysis on the Seurat guidelines^23^ and set the boundaries for the main parameters (genes expressed per cell and mitochondrial content) on quality control, as included in the related Github repository. We retained all cells expressing more than 750 genes and less than 5000, with a mitochondrial faction lower than 0.15. We normalized the data and extracted the variable features, then scaled the data regressing against the fraction of the mitochondrial content. We ran a PCA on the scaled data, and used the first 25 components to calculate the UMAP projection and the neighboring cells. Clusters were extracted with the Louvain algorithm (resolution = 0.1).

**Differential gene expression** was calculated with a Poisson model, minimal fraction of cells expressing the gene = 0.25 and module of the base-2 logarithm of the fold change, |log2FC| > 0.5. For CBD signatures, we considered as DEGs the genes showing |log2FC| > 1, i.e., an expression at least doubled or halved when comparing conditions, both globally (i.e., considering the merged data set) and in at least two out of three individuals (i.e., considering cells from a single individual).

### Inflammation score

We used the Seurat function AddModuleScore to calculate the inflammation score. We based the score on the genes listed in the GO term *positive regulation of inflammatory response* (GO:0050729), considering genes that are expressed in our data (101 genes total). Briefly, this score is based on the average expression levels of the GO genes, subtracted by the aggregated expression of a control set of 100 randomly selected genes. To make the expression levels comparable, genes are binned based on averaged expression (24 bins).

### STRING database analysis

We used our six myeloid cell DEGs as the input for a STRING (v. 11.5)^38^ analysis. To calculate the p-value, we proceeded empirically, as follows: We considered all the human protein-protein interactions of STRING (∼20 thousand proteins involved in ∼12 billion interactions); we iteratively extracted 6 random genes, and counted how many interactions were present in the subset with a score ≥ 0.9, the threshold of our analysis. If the random subset had 4 or more interactions (our signature has 4), we counted a hit. We retrieved 5 hits over 50,000 iterations. Pathway analysis was conducted with STRING. First DEG interactors where added with highest confidence (0.9) and allowing up to 50 interactions. All p-values are FDR-adjusted.

**Cell-cell communication variation** was inferred with CellChat^42^. Plasma cells were excluded by the analysis because of the small size of the cell population. The cell-cell communication database we used is CellChatDB based on Human cells. The population size parameter was set to TRUE in the computeCommunProb function as suggested by the manual for non-sorted cells. Other parameters were used at default value.

## Data Availability statement

The code to obtain our results is available on github (https://github.com/smarini/CBD_HIV_scRNAseq). Our data are available on GEO (https://www.ncbi.nlm.nih.gov/geo/query/acc.cgi?acc=GSE228652).

## Author contributions

Conception/design of work: SM, RLC, PB, CNM; Acquisition of funding: SM, CNM; Methods/analysis/interpretation of data: SM, AH, MNC, MS, CNM; Writing the first draft: SM, MNC; Editing: AH, CNM; All authors read and approved the final manuscript.

